# Evaluation of 2022 Nursing Intake Forms from Massachusetts Jails: Content Analysis and Accuracy Assessment

**DOI:** 10.1101/2023.10.24.23297482

**Authors:** Emily D. Grussing, Bridget Pickard, Emma Smyth, Lauren Brinkley-Rubinstein, Alysse G. Wurcel

## Abstract

The medical intake process is usually the first clinical interaction that people have after being incarcerated. In many states, the first vital person conducting this assessment is a nurse. In this project, nursing intake forms from Massachusetts jails were analyzed for the offer of 15 different screening and preventative services (e.g., HIV testing). Then, Health Services Administrators (HSAs) at the jails were asked to review and comment on the assessment. The average agreement rate between the project team members and HSAs was 73%. Twelve of the 14 jails had questions about history of HIV or HCV on the nursing intake form. Only 5 facilities discussed PrEP. There is potential to standardize the nursing intake form and therefore medical intake process. This must be done with key stakeholder groups, in particular nurses, to best understand the logistics of the complex medical intake process.

## Introduction

As the source of healthcare for over one million adults in the United States every year, the systems of healthcare delivery in jails should be routinely evaluated for areas of improvement (Carson, 2022). Nurses and nurse-administrations are integral to and at the center of these healthcare systems.

People who are detained and incarcerated in criminal-legal settings face barriers to quality healthcare and have higher rates of infectious diseases, mental illness, and cardiovascular disease (Ahalt et al., 2013; Camplain et al., 2021; Dumont, 2013; Dumont et al., 2012; Freudenberg & Heller, 2016; Johnson et al., 2013; Massoglia et al., 2014). In order to facilitate continuity of care and offer preventative care, there is a medical intake process in most carceral settings where the person who is being detained interacts with a nurse or other member of the jail staff to review medical history, active medical treatments, and screen for psychiatric illness (Anno, 2001; Maner et al., 2022; Ndeffo-Mbah et al., 2018). This information is collected using a questionnaire, or medical intake form. Medical intake forms are completed almost universally across U.S. jails, prisons, and detention centers, although there is variability about who asks the questions and what questions are asked (Carda-Auten et al., 2022; Maner et al., 2022). Beyond asking about medical history, the CDC Infectious Diseases Corrections 2022 guidelines recommend that medical intake form should include offering services, including evaluation for HIV, hepatitis, tuberculosis, gonorrhea, chlamydia, and syphilis (CDC, 2022b). The same guidelines also advise that vaccines and PrEP should be offered (CDC, 2022b).

In Massachusetts, the initial medical intake screening is always completed by a nurse, so this document is henceforth referred to as the nursing intake form for the purpose of this study. The same study team published an evaluation of 2018 nursing intake forms in Massachusetts and found heterogeneity in the infectious diseases questions asked during nursing intake (Wurcel et al., 2021). About 85% of jails asked about a history of HIV, but only 50% offered HIV testing (Wurcel et al., 2021). Half of the jails asked about previous HCV infection, and only 2 offered testing (Wurcel et al., 2021). Since the study team’s 2018 evaluation, the COVID19 pandemic brought increased attention to the process of COVID19 screening for people entering jail as well as offering COVID19 vaccination.

Prison and jail scholars interested in learning more about medical intake process have requested jail medical intake forms via the Freedom of Information Act (FOIA) (Yaremko, 2020). There is an increase in research reports analyzing the jail medical intake forms as a marker of healthcare. However, the accuracy of these content analyses has not been reviewed by the most critical jail employees: nurses and nurse-administrators.

## Objective

The goal of this evaluation was to perform a content analysis on Massachusetts nursing intake forms from 2022. To complete the summative content analysis, researchers collected nursing intake forms and then had health services administrators (HSAs) from every jail support or refute the researchers’ interpretation of data on the nursing intake forms, allowing researchers to revise findings if indicated.

## Methods

### Setting

Massachusetts has 14 counties and 15 jails. All the Massachusetts jails have accreditation from the American Correctional Association, the National Commission on Correctional Health Care, or both. The jails either have healthcare provided by the county, a for-profit medical organization, or a hybrid model. One county jail was excluded from this analysis due it is extremely small size and unique healthcare delivery system.

### Design

This is a descriptive qualitative study design with summative content analysis.

### Sample

The research team collected nursing intake forms from a convenience sample of fourteen jails in Massachusetts. HSAs are typically nurse-administrators who oversee the healthcare provided at jails. The nursing checklists were requested via email twice, and we followed up with one phone call.

### Research team checklist development

The project team modified the previous evaluation research checklist on nursing intake from 2018 (Wurcel et al., 2021) to evaluate the nursing intake forms again in 2022 (Supplement 1). The modifications included surveying for the presence of questions on the intake form that addressed LGBTQIA+ status, offering COVID19 vaccination/history of vaccination, history of COVID19 infection, Sickle Cell disease, and offering STD testing. These modifications stemmed from an interest in taking a more holistic, updated view of infections and chronic diseases that affect infections that should be screened at intake. In particular, since the 2018 study, there have been initiatives in healthcare and jail settings to ask about LGBTQA+ status (Bacak et al., 2018). Similar to previous projects, tuberculosis screening was not included in the assessment. The project team made this decision because tuberculosis screening in Massachusetts is universally implemented across jails in line with 2006 CDC recommendations and national accreditation requirements (CDC, 2006; NCCHC, 2023).

#### Research team content analysis

Two Project teams members, BP and ES, analyzed nursing intake forms for items on the check list topics. The team members would answer “Yes,” “No,” or “Unsure.” Then two team members’ findings were compared, and any differing findings were resolved through repeat examination of the form and involvement of a third research team member to tie-break, if necessary. One hundred percent inter-rater reliability was achieved.

### Agreement check survey development

Based on the results of the team members’ analysis (see research team survey development), a customized electronic survey was sent to each HSA at every facility with the findings of their facility’s nursing intake form. An example of a survey is included as a supplement (Supplement 2). The HSAs were asked to: (1) agree with the evaluation; or (2) disagree with the evaluation. If the HSA disagreed, they were asked to explain in a free text box why they disagreed with the findings.

### Expert review incorporation

The research team recorded the responses given by the HSAs. If an HSA stated a topic was addressed, the research team would consider that topic as addressed during nursing intake. If an HSA stated a topic was not addressed, the research team would consider that topic as not addressed during intake. The research team would also record that that topic was changed from yes to no or no to yes after expert review by the HSA. The research team also collected the HSA’s free text reasons for why they disagreed with the research team’s content analysis findings.

### Ethics statement

The collection of nursing intake forms was exempt as non-human subjects’ research. The nursing intake forms collected for analysis were blank, so they contained no identifying information. The survey of the HSAs was granted as exempt from consent by the Tufts University Health Sciences Institutional Review Board (IRB).

## Results

All 14 Massachusetts jails sent their nursing intake forms and responses to the surveys. The jails were anonymized as Jails A-N. Of note, two jails only completed page one of the survey (five questions). All other jails completed 15/15 questions. The results of the initial content analysis are displayed in Figure 1. Twelve of the 14 jails had questions about history of HIV or HCV on the nursing intake form. The least commonly covered topic was offering PrEP, with only five facilities discussing PrEP during nursing intake. The average agreement rate between the team members and HSAs was 73%. Free text responses fell into six broad categories: (1) topic is covered elsewhere during nursing intake (unspecified); (2) topic is covered elsewhere during nursing intake (specified); (3) topic is addressed by a broader question during nursing intake; (4) test is mandated for everyone, so not asked during nursing intake; (5) topic is addressed, but verbally; and (6) topic has been added since documents were shared with project team. These results can be found in Table 1. The most common reason for changing a response was that the topic is covered during an unspecified part of the nursing intake, with 12 responses falling into this category.

**Figure 1.**
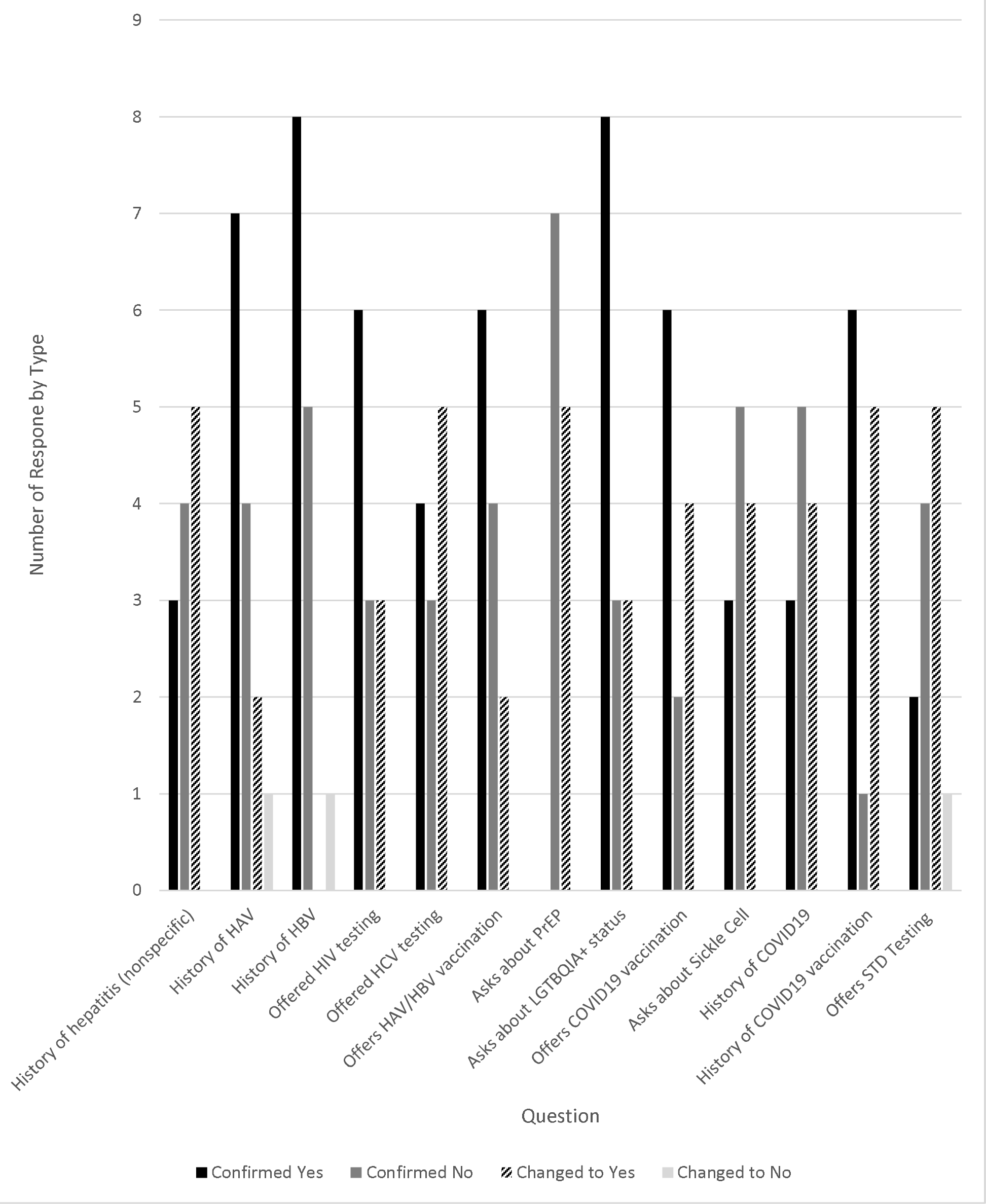
Responses by question.

**Table 1.** Classification of Free Text Responses When HSA disagreed with Researcher Content Analysis.

|  |  |
| --- | --- |
| Topic is covered elsewhere during intake (unspecified) | 12 |
| Topic is covered elsewhere during intake (specified) | 10 |
| Topic is addressed by a broader question during nursing intake | 8 |
| Test is mandated for everyone, so not asked in nursing intake | 3 |
| Topic is addressed, but verbally | 3 |
| Topic has been added since document was shared | 2 |

## Discussion

Although the nursing intake form may seem like a straightforward, accessible method for researchers, policy experts, and advocates to analyze the healthcare topics discussed during medical intake at jails, this study showed that the nursing intake form did not consistently reflect healthcare delivery. There are several reasons why this finding is an important addition to the literature. First, researchers, public health experts, and litigators may use nursing intake forms to evaluate the care provided at jails. Based on our analysis, the nursing intake forms do not capture all the types of healthcare services provided. Second, a goal of nursing intakes forms is to operationalize best practices so that the clinical evaluation can be equitable. Several jails reported that the infectious disease testing and vaccination was offered, just not at the time of nursing intake or on a separate form from the nursing intake form. The fragmentation of infectious diseases testing and vaccination into different systems and clinical interactions creates opportunities for heterogeneity in who gets offered or is able to access care. Third, despite being in a small state, there were differences in how jails incorporated infectious diseases testing and vaccination into the nursing intake form in Massachusetts. These differences could be exponentially greater in larger states.

The *Guidelines for the Management of an Adequate Delivery System* published by the National Commission of Correctional Health Care describes the medical intake process as the most important standard for correctional facilities to meet (Anno, 2001). The nursing intake form in Massachusetts is the first step in a multi-layered process of acute and chronic healthcare including organizing emergency triage; coordinating continuity of care; assessing current health status including mental and dental health; and offering a variety of optional preventative services (e.g., screenings and vaccination) (Anno, 2001). The rate of deaths due to alcohol in United States jails doubled between 2001-2018 (Carson 2021), and two-thirds of people who are incarcerated meet standards for substance use disorder/dependence (Bronson, 2017). The mental and physical status of a person who is newly incarcerated may be impaired because of acute intoxication or withdrawal, leading to incorrect answers or answers to questions that may not represent their true wishes. Information about medical care does not pass easily from the jail to community or back (Butler, 2014). All of these factors complicate the process of working towards continuity of care for infectious diseases and providing the highest level of evidence-based interventions.

The creation of a universal infectious diseases intake form for jails that represents best practices is a feasible next step to encourage uniform application of best public health practices. Despite several obstacles, tuberculosis screening has been implemented across the U.S. in jails and prisons, with incredible impact in reduction of tuberculosis transmission both in the carceral setting and in the community (CDC, 2022a; Puisis et al., 1996; Saunders et al., 2001; Stewart et al., 2022). The creation of a universal infectious diseases medical intake form will require getting feedback from key stakeholders across the criminal-legal spectrum on the best time to ask questions, the way to ask questions, and the potential downstream consequences, such as increased medical costs, that need to be considered.

As the first people who meet and triage people who are incarcerated, nurses best understand the challenges of accessing accurate health information at intake into a carceral facility. Nurse-administrators understand the procedures for standardizing healthcare practices and assessing their efficacy. People who are incarcerated understand the lived experience of being detained and can provide expert first-hand feedback on the intake process, including what services they think they should be offered and what questions should be asked of and answered for them. As evidenced by this current review, there are lapses in questions asked and services offered during the intake process. The key to improving any care given during intake to a carceral facility is asking nurses and nurse-administrators on how to best do it.

Limitations of this report include focusing on a single state and getting feedback from only one person working at the jails. Due to the substantial differences that exist between state-run jails, a similar analysis in a different state may yield different findings. Some states do not have intake forms asked by nurses, which may require investigators in other states to modify their assessment of the medical intake forms. This study was only conducted in jails, and the medical intake process in prisons may be different. It should be noted that two jails only completed five questions of our 15-question electronic survey. We believe this was because the survey was three pages long, with five questions per page, and respondents may have not seen the arrow at the bottom of the screen directing them to the rest of the survey. The survey questions sent to each facility were in a different order, so the 10 questions that were not answered were different for each facility. Therefore, the study team believes the missing questions did not greatly impact study results, as they only limited responses per question by one or two at most.

Despite these limitations, this work adds to the growing body of literature analyzing jail medical intake forms with goal of improving jail healthcare. It also highlights the shortcomings of these analyses. This work also calls for and outlines the potential process for standardizing the medical intake process, particularly by incorporating a consistent infectious diseases intake form, which would need to be developed with nursing feedback, which could help reduce disparity and promote best public health practices.

## Data Availability

All data produced in the present study are available upon reasonable request to the authors.

## References

Ahalt, C., Trestman, R. L., Rich, J. D., Greifinger, R. B., & Williams, B. A. (2013). Paying the price: the pressing need for quality, cost, and outcomes data to improve correctional health care for older prisoners. J Am Geriatr Soc, 61(11), 2013–2019. 10.1111/jgs.12510

Anno, B. J. (2001). Guidelines for the Management of an Adequate Delivery System (Vol. 2021). Natioanl Commission on Correctional Health Care.

Bacak, V., Thurman, K., Eyer, K., Qureshi, R., Bird, J. D. P., Rivera, L. M., & Kim, S. A. (2018). Incarceration as a Health Determinant for Sexual Orientation and Gender Minority Persons. Am J Public Health, 108(8), 994–999. 10.2105/AJPH.2018.304500

Bronson, J. S., Jessica. (2017). Drug Use, Dependence, and Abuse Among State Prisoners and Jail Inmates, 2007-2009 (Special Report, Issue. B. o. J. Statistics. https://bjs.ojp.gov/content/pub/pdf/dudaspji0709.pdf

Butler, B. (2014). Health information exchange between jails and their communities: a bridge that is needed under healthcare reform. Perspect Health Inf Manag, 11(Winter), 1b. https://www.ncbi.nlm.nih.gov/pubmed/24808809

Camplain, R., Lininger, M. R., Baldwin, J. A., & Trotter, R. T., 2nd. (2021). Cardiovascular Risk Factors among Individuals Incarcerated in an Arizona County Jail. Int J Environ Res Public Health, 18(13). 10.3390/ijerph18137007

Carda-Auten, J., Dirosa, E. A., Grodensky, C., Nowotny, K. M., Brinkley-Rubinstein, L., Travers, D., Brown, M., Bradley-Bull, S., Blue, C., & Rosen, D. L. (2022). Jail Health Care in the Southeastern United States From Entry to Release. Milbank Q, 100(3), 722–760. 10.1111/1468-0009.12569

Carson, E. A. (2022). Prisoners in 2021 – Statistical Tables. U.S. Department of Justice. Retrieved April 20, 2023 from chrome-extension://efaidnbmnnnibpcajpcglclefindmkaj/https://bjs.ojp.gov/sites/g/files/xyckuh236/files/media/document/p21st.pdf

CDC. (2006). Slide Set — Tuberculosis (TB) in Correctional Settings: What Corrections Staff Need to Know. Retrieved March 20 from chrome-extension://efaidnbmnnnibpcajpcglclefindmkaj/https://www.cdc.gov/tb/publications/slidesets/correctionsalstaff/correctional-textonlyversion.pdf

CDC. (2022a). Correctional Facilities. CDC. Retrieved March 20, 2023 from https://www.cdc.gov/tb/topic/populations/correctional/default.htm

CDC. (2022b). Testing, Vaccination, and Treatment for HIV, Viral Hepatitis, TB, and STIs (Correction Health: Recommendations and Guidance, Issue. https://www.cdc.gov/correctionalhealth/rec-guide.html

Dumont, D. M., Allen, S., Brockman, B., Alexander, N.E., Rich, J.D.. (2013). Incarceration, Community Health, and Racial Disparities. Journal of Health Care for the Poor and Underserved, 21, 78–88, Article 1. 10.1353/hpu.2013.0000

Dumont, D. M., Brockmann, B., Dickman, S., Alexander, N., & Rich, J. D. (2012). Public health and the epidemic of incarceration. Annu Rev Public Health, 33, 325–339. 10.1146/annurev-publhealth-031811-124614

Freudenberg, N., & Heller, D. (2016). A Review of Opportunities to Improve the Health of People Involved in the Criminal Justice System in the United States. Annu Rev Public Health, 37, 313–333. 10.1146/annurev-publhealth-032315-021420

Johnson, A. P., Macgowan, R. J., Eldridge, G. D., Morrow, K. M., Sosman, J., Zack, B., Margolis, A., & Project, S. S. G. (2013). Cost and threshold analysis of an HIV/STI/hepatitis prevention intervention for young men leaving prison: Project START. AIDS Behav, 17(8), 2676–2684. 10.1007/s10461-011-0096-7

Maner, M., Omori, M., Brinkley-Rubinstein, L., Beckwith, C. G., & Nowotny, K. (2022). Infectious disease surveillance in U.S. jails: Findings from a national survey. PLoS One, 17(8), e0272374. 10.1371/journal.pone.0272374

Massoglia, M., Pare, P. P., Schnittker, J., & Gagnon, A. (2014). The relationship between incarceration and premature adult mortality: gender specific evidence. Soc Sci Res, 46, 142–154. 10.1016/j.ssresearch.2014.03.002

NCCHC. (2023). Health Assessment. NCCHC. Retrieved March 20, 2023 from https://www.ncchc.org/q-a/health-assessment/

Ndeffo-Mbah, M. L., Vigliotti, V. S., Skrip, L. A., Dolan, K., & Galvani, A. P. (2018). Dynamic Models of Infectious Disease Transmission in Prisons and the General Population. Epidemiol Rev, 40(1), 40–57. 10.1093/epirev/mxx014

Puisis, M., Feinglass, J., Lidow, E., & Mansour, M. (1996). Radiographic screening for tuberculosis in a large urban county jail. Public Health Rep, 111(4), 330–334. https://www.ncbi.nlm.nih.gov/pubmed/8711100

Saunders, D. L., Olive, D. M., Wallace, S. B., Lacy, D., Leyba, R., & Kendig, N. E. (2001). Tuberculosis screening in the federal prison system: an opportunity to treat and prevent tuberculosis in foreign-born populations. Public Health Rep, 116(3), 210–218. 10.1093/phr/116.3.210

Stewart, R. J., Raz, K. M., Burns, S. P., Kammerer, J. S., Haddad, M. B., Silk, B. J., & Wortham, J. M. (2022). Tuberculosis Outbreaks in State Prisons, United States, 2011-2019. Am J Public Health, 112(8), 1170–1179. 10.2105/AJPH.2022.306864

Wurcel, A. G., Chen, G., Zubiago, J. A., Reyes, J., & Nowotny, K. M. (2021). Heterogeneity in Jail Nursing Medical Intake Forms: A Content Analysis. J Correct Health Care, 27(4), 265–271. 10.1089/jchc.20.04.0018

Yaremko, J. W., Kevin. (2020). Use of Freedom of Information Requests in Prison and Jail Studies. Center for Access to Information and Justice. chrome-extension://efaidnbmnnnibpcajpcglclefindmkaj/https://www.uwinnipeg.ca/caij/docs/reports/use-of-freedom-of-information-requests-in-prison-and-jail-studies.pdf

Yehia, B. R., Schranz, A. J., Umscheid, C. A., & Lo Re, V., 3rd. (2014). The treatment cascade for chronic hepatitis C virus infection in the United States: a systematic review and meta-analysis. PLoS One, 9(7), e101554. 10.1371/journal.pone.0101554

