## Supplement 1 for "Evaluation of 2022 Nursing Intake Forms from Massachusetts Jails: Content Analysis and Accuracy Assessment"

|  | Jail A | Jail B | Jail C | Jail D | Jail E | Jail F | Jail G | Jail H | Jail I | Jail J | Jail K | Jail L | Jail M | Jail N |
| --- | --- | --- | --- | --- | --- | --- | --- | --- | --- | --- | --- | --- | --- | --- |
| History of HIV/previous HIV testing asked?* |  |  |  |  |  |  |  |  |  |  |  |  |  |  |
| History of HCV/previous HCV testing asked? |  |  |  |  |  |  |  |  |  |  |  |  |  |  |
| History of hepatitis (nonspecific) asked? |  |  |  |  |  |  |  |  |  |  |  |  |  |  |
| History of HAV asked? |  |  |  |  |  |  |  |  |  |  |  |  |  |  |
| History of HBV asked? |  |  |  |  |  |  |  |  |  |  |  |  |  |  |
| Offers HIV testing? |  |  |  |  |  |  |  |  |  |  |  |  |  |  |
| Offers HCV testing? |  |  |  |  |  |  |  |  |  |  |  |  |  |  |
| Offers HAV/HBV vaccination? |  |  |  |  |  |  |  |  |  |  |  |  |  |  |
| Asks about PrEP? |  |  |  |  |  |  |  |  |  |  |  |  |  |  |
| Asks about LGTBQIA+ status? |  |  |  |  |  |  |  |  |  |  |  |  |  |  |
| Offers COVID19 vaccination? |  |  |  |  |  |  |  |  |  |  |  |  |  |  |
| Asks about Sickle Cell? |  |  |  |  |  |  |  |  |  |  |  |  |  |  |
| History of COVID19 asked? |  |  |  |  |  |  |  |  |  |  |  |  |  |  |
| History of COVID19 vaccination asked? |  |  |  |  |  |  |  |  |  |  |  |  |  |  |
| Offers STD Testing? |  |  |  |  |  |  |  |  |  |  |  |  |  |  |

*For all questions for each jail, research assistants answered yes or no.

Supplement 1. Checklist of questions
