## Supplement 2 for "Evaluation of 2022 Nursing Intake Forms from Massachusetts Jails: Content Analysis and Accuracy Assessment"

**Consent Information**

Thank you for your interest in helping us double-check our work!

This research survey is voluntary, anonymous, and you can choose to stop participation at any time without penalty. You are being invited to take part in this survey because you are a health service administrator at a Massachusetts jail. We have done our best to collect this information correctly, but we realize that some jails, for example, may oﬀer HIV testing in a separate form.

Data from this survey will be analyzed by Dr. Alysse Wurcel’s research team at Tufts Medical Center. The survey should take about 15 minutes to complete, and there will be no payment/incentive/beneﬁt for your participation. There are no foreseeable risks if you choose to complete this survey as we are not collecting any conﬁdential or identiﬁable information from you. The alternative to participating in this study is to not participate.

If you would like to be contacted for further research opportunities, or have any comments or concerns, please email Alysse Wurcel. If you have questions about your rights as a research study subject, call the Tufts University Health Sciences Institutional Review Board (IRB) at (617) 636-7512. This study has been reviewed by the Tufts University Health Sciences IRB.

In your nursing medical intake forms, we found:

A question about the individual’s HIV history/previous HIV testing.

Do you agree that [REDACTED] jail asks the individual about their HIV history/previous HIV testing upon intake?

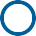
 Yes

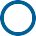
 I don't know
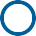
 No

A question about the individual’s HCV history/previous HCV testing.

Do you agree that [REDACTED] jail asks the individual about their HCV history/previous HCV testing upon intake?

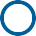
 Yes

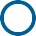
 I don't know
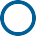
 No

A question asking about the individual’s history of HAV.

Do you agree that [REDACTED] jail asks the individual about their history of HAV upon intake?

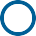
 Yes

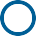
 I don't know
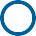
 No

A question asking about the individual’s history of HBV.

Do you agree that [REDACTED] jail asks the individual about their history of HBV upon intake?

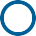
 Yes

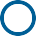
 I don't know
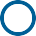
 No

A question asking about the individual’s LGBTQIA+ status.

Do you agree that [REDACTED] jail asks the individual about their LGBTQIA+ status upon intake?

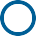
 Yes

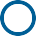
 I don't know
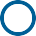
 No

In your nursing medical intake forms, we did not ﬁnd:

A question asking about the individual’s history of hepatitis (nonspeciﬁc).

Do you agree that [REDACTED] jail does not ask about the individual’s history of hepatitis (nonspeciﬁc) upon intake?

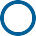
 Yes

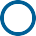
 I don't know

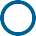
 No (if no, please indicate the way in which you ask this information in the text box below)

That [REDACTED] jail oﬀers HIV testing.

Do you agree that [REDACTED] jail does not oﬀer HIV testing upon intake?

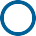
 Yes

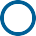
 I don't know

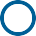
 No (if no, please indicate the way in which you ask this information in the text box below)

That [REDACTED] jail oﬀers HCV testing.

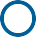
Do you agree that [REDACTED] jail does not oﬀer HCV testing upon intake?

That [REDACTED] jail oﬀers HAV/HBV vaccination.

Do you agree that [REDACTED] jail does not oﬀer HAV/HBV vaccination upon intake?

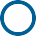
 Yes

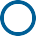
 I don't know

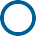
 No (if no, please indicate the way in which you ask this information in the text box below)

A question asking the individual about PrEP.

Do you agree that [REDACTED] jail does not ask about PrEP upon intake?

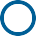
 Yes

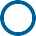
 I don't know

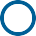
 No (if no, please indicate the way in which you ask this information in the text box below)

That [REDACTED] jail oﬀers COVID-19 vaccination.

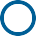
Do you agree that [REDACTED] jail does not oﬀer COVID-19 vaccination upon intake?

A question asking the individual about Sickle Cell.

Do you agree that [REDACTED] jail does not ask the individual about Sickle Cell upon intake?

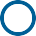
 Yes

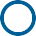
 I don't know

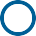
 No (if no, please indicate the way in which you ask this information in the text box below)

A question asking the individual about their history of COVID-19.

Do you agree that [REDACTED] jail does not ask the individual about their history of COVID-19 upon intake?

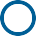
 Yes

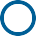
 I don't know

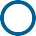
 No (if no, please indicate the way in which you ask this information in the text box below)

A question asking the individual about their history of COVID-19 vaccination.

Do you agree that [REDACTED] jail does not ask the individual about their history of COVID-19 vaccination upon intake?

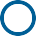
 Yes

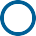
 I don't know

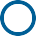
 No (if no, please indicate the way in which you ask this information in the text box below)

That [REDACTED] jail oﬀers STD testing.

Do you agree that [REDACTED] jail does not oﬀer STD testing upon intake?

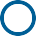
 Yes

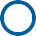
 I don't know

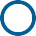
 No (if no, please indicate the way in which you ask this information in the text box below)

[Powered by Qualtrics](https://www.qualtrics.com/powered-by-qualtrics/?utm_source=internal%2Binitiatives&utm_medium=survey%2Bpowered%2Bby%2Bqualtrics&utm_content=%7B~BrandID~%7D&utm_survey_id=%7B~SurveyID~%7D)
